# Impact of the COVID-19 pandemic on the uptake of intermittent sulphadoxine-pyrimethamine for prevention of malaria amongst pregnant women in Sierra Leone

**DOI:** 10.1101/2025.01.21.25320891

**Authors:** William Tilima Pessima, Bailah Molleh, Musa Silla-Kanu, Brenda Stafford, Wani Kumba Lahai, Abdul Mac Falama, Sharmistha Mishra, Adrienne K. Chan, Stephen Sevalie, Sulaiman Lakoh, Michael Lahai

## Abstract

**Background:** Malaria in pregnancy contributes to high maternal mortality, warranting the adoption of intermittent preventive treatment (IPTp) of malaria in pregnancy in Sierra Leone in 2017. Since then, the uptake of IPTp has increased but it is unclear whether COVID-19 has any impact on these services. We aimed to assess the impact of COVID-19 on the uptake of IPTp among pregnant women in Sierra leone at district, regional, and national levels.

**Methods:** A retrospective cross-sectional study design was used to assess the monthly aggregated data from DHIS2 for all 5 regions of Sierra Leone. We describe the trends of IPTp1, IPTp2, and IPTp3+ uptake at national, regional, and district levels before, during and after COVID-19. Data was analyses using STATA. The median and interquartile range for each outcome in each period were calculated.

**Results:** The median monthly uptake of total IPTp was the lowest before COVID (53,244) but increased during COVID-19 (55,257) and after COVID-19 (59,439). IPTp1 uptake was lowest during COVID-19 (20,338) but highest after COVID-19 (20,914). All three variables (IPTp1, IPTp2 and IPTp3) showed a clear decline in uptake in early 2020 and recovered within 3 months in a similar pattern of magnitude, except in Eastern Sierra Leone. Both IPTp2 and IPTp3 showed more pronounced increases in uptake after recovery than IPTp1 and they both experienced sustained increases in the post-COVID-19 period.

**Conclusion:** The negative impact of the COVID-19 pandemic in reducing IPTp uptake was followed by rapid recovery, underscoring the resilience of the Sierra Leone’s health system in response to public health emergencies. However, the delay in recovery in the IPTp uptake in the Eastern Sierra Leone highlight the need for strong differentiated regional strategies in protecting essential health services during public health emergencies.

## Introduction

Malaria is responsible for 822,000 children with low birth weight in sub-Saharan Africa, more than half of which occur in the West African region (1). About 6% of malaria deaths in sub-Saharan Africa are due to low birth weight (2). Historically, low birth weight has been a major problem in Sierra Leone, caused mostly by infections such as malaria (3). Malaria in pregnancy is associated with a range of complications, including low birth weight, maternal hypoglycemia, intrauterine growth retardation, placental malaria, fetal hypotrophy, miscarriage, abortion, anaemia and preterm delivery, which explains the contribution of malaria to higher maternal and neonatal mortality rates (4). A meta-analysis of studies primarily from sub-Saharan Africa showed that successful implementation of malaria and other interventions could reduce the risk of anaemia and child mortality (5).

Intermittent Preventive Treatment for malaria in pregnancy (IPTp) with Sulfadoxine-Pyrimethamine (SP) is an evidence-based programmatic intervention that can prevent the adverse outcomes of malaria in pregnancy (1). Strong programmatic efforts by national and international agencies, supported by political commitment, are needed to achieve substantial progress in the uptake of IPTp-SP and, in turn, reduce the prevalence of malaria in pregnancy (6). Over the past decade, most sub-Saharan African countries, including Sierra Leone, have introduced IPTp policies for pregnant women. The current policy of the Sierra Leone malaria programme is to administer IPTp-SP at 13 weeks of gestation since 2017 and every four weeks thereafter. Similar policies had been implemented in other African countries (7) (8). Similar policies had been implemented in other African countries (7) (8).

In 2020, the early phase of the COVID-19 pandemic resulted in health system adjustments with the need for reallocation of the already limited resources away from the national malaria control of resources to support surveillance and other healthcare services. Concerns about the association of COVID-19 with hospitals or hospital visits and the closure of some health facilities to provide services to COVID-19 patients, have changed people’s health-seeking behaviour and are among the barriers to access services including, IPTp-SP (9). These events led to disruptions in hospital care and significantly affected access to IPTp-uptake for pregnant women in Sierra Leone. Several previous studies on IPTp uptake have focused on the prevalence and predictors of IPTp uptake among pregnant women (10).

Existing research evaluating the uptake of IPTp in Africa and Sierra Leone was more focused on the factors affecting IPTp-uptake with scarce research information on IPTp disaggregated data by region and district and this was not assessed in the recent multi-cluster indicator survey in Sierra Leone (11). Our study aims to assess the impact of COVID-19 on the uptake of Intermittent Preventive Therapy (IPTp) for malaria in pregnancy in Sierra Leone at the district, regional, and national levels to inform national policy around the resiliency of the malaria control programming.

## Methods

### Study Design

We conducted a retrospective cross-sectional study using monthly aggregated secondary data on IPTp uptake among pregnant women who access ANC services in their 2^nd^ and 3^rd^ trimester of pregnancy extracted from the District Health Information system-2 (DHIS2) platform for all the 5 regions of Sierra Leone.

### Study Setting

The health system is divided into three tiers: primary health care and extended community health programs, secondary health care, and tertiary health care. The primary healthcare system has 1430 peripheral health units (PHUs). IPTp is given to pregnant women during routine ANC in PHUs and by community health workers. The current national IPTp policy for IPTp recommends giving SP at 13 weeks of pregnancy with subsequent doses every 4 weeks. We used data on IPTp uptake among pregnant women from all 5 regions in Sierra Leone: Eastern (Kailahun, Kenema, and Kono districts), Western Area (Urban and Rural), Northern (Bombali, Falaba, Koinadugu, and Tonkolili districts), Northwest (Kambia, Karene, and Port Loko districts), and the Southern region (Bo, Bonthe, Moyamba, and Pujehun districts).

### Data Sources and Variables

Aggregated monthly data on IPTp uptake was extracted from the DHIS-2 platform, a national health information system on September 4, 2023. The data were initially collected from individual patients in the PHUs, aggregated monthly, and then forwarded to the District Monitoring and Evaluation Officers for review and entry into the DHIS-2 platform. Each dose is for one eligible person each month. IPTp1 is the first dose of IPTp given in the 2^nd^ or 3^rd^ trimester, IPTp2 is the second dose of IPTp given 4 weeks after the first dose, IPTp3+ is the third dose of IPTp given 4 weeks after the second dose, and any subsequent doses given up until delivery (including the 4^th^ to 7^th^ doses).

Monthly data from January 2019-December 2023 was extracted to capture seasonal variation in service utilization and to reflect IPTp uptake in three time periods: January 2019-Feb 2020 (before COVID-19; March 2020-Feb 2022 (during COVID-19); March 2022-December 2022 (after COVID-19). Variables extracted included: the number of pregnant women who received IPTp (IPTp1, IPTp2, IPTp3+) per month in the second (gestational weeks 13 to 24) and third (gestational weeks 24 to delivery) trimesters for 16 districts (divided into five regions).

### Data Management and Analysis

The data in Excel format were cleaned, coded, and validated. Analyses and data visualization were performed in STATA (version 15.1 Stata Corp LLC, College Station, TX). We describe and visualize trends over time to demonstrate national and regional variation in IPTp1, IPTp2, and IPTp3+ uptake before, during, and after COVID-19. The median and interquartile range for each outcome in each period was calculated for descriptive statistics.

### Ethical consideration

Ethics approval was obtained from the Sierra Leone Ethics and Scientific Review Committee (SLESRC number: 019/05/2023). Since routinely aggregated data, informed consent did not apply with approval for data extraction and use by the Chief Medical Officer, Ministry of Health, Sierra Leone. Finally, the authors did not have access to information that could identify individual participants during or after data collection.

## Results

### Uptake of IPTp before, during and after COVID-19

Data from 14 months of national IPTp uptake before COVID-19, 24 months during COVID-19, and 10 months after COVID-19 were extracted from the DHIS-2. The median monthly uptake of total IPTp was the lowest before COVID-19 (53,244) but increased during COVID-19 (55,257). The exception was the median monthly uptake of IPTp1, which was lowest during COVID-19 (20,338), but highest after COVID-19 (20,914) (***Table 1)***.

**Table 1.** Summary of IPT uptake over three time periods in Sierra Leone.

|  | Pre-COVID |  | During COVID |  | Post-COVID |  |
| --- | --- | --- | --- | --- | --- | --- |
|  | Mar 2019 to Feb 2020 |  | Mar 2020 to Feb 2022 |  | Mar 2022 to Dec 2022 |  |
|  | 12 months |  | 24 months |  | 10 months |  |
|  | Median | (IQR) | Median | (IQR) | Median | (IQR) |
| **Uptake per month, National level** |  |  |  |  |  |  |
| Total (IPTp-1, IPTp-2, IPTp-3+) | 53,244 | (52,344; 53,662) | 55,257 | (52,908; 57,236) | 59,439 | (58,670; 61,288) |
| IPTp-1 | 20,405 | (19,491; 21,280) | 20,338 | (19,330; 20,901) | 20,914 | (20,362; 21,914) |
| IPTp-2 | 17,757 | (16,943; 18,103) | 17,859 | (16,882; 18,725) | 19,758 | (18,859; 20,556) |
| IPTp-3+ | 14,597 | (13,884; 15,093) | 16,982 | (15,756; 17,861) | 18,638 | (18,513; 19,186) |

IPTp (intermittent preventive therapy in pregnancy), IPTp-3+ refers to the 3<sup>rd</sup> dose and beyond). IQR (inter-quartile range).

### Differential uptake of IPTp1, IPTp2, and IPTp3+ before, during and after COVID-19

During these three time periods, there were small fluctuations in the uptake of all forms of IPTp (total IPTp, IPTp1, IPTp2 and IPTp3+). Before COVID-19, the IPTp was stable with a decline in all forms of IPTp uptake in the last three months and the start of the COVID-19 period, specifically from January-March 2020. The uptake of all forms of IPTp recovered in the first three months of the COVID-19 period (until June 2020), with a lower magnitude decline and recovery over a couple of months before steadily increasing during late COVID-19 and after COVID-19 (**Figure 1)**.

**Figure 1:**
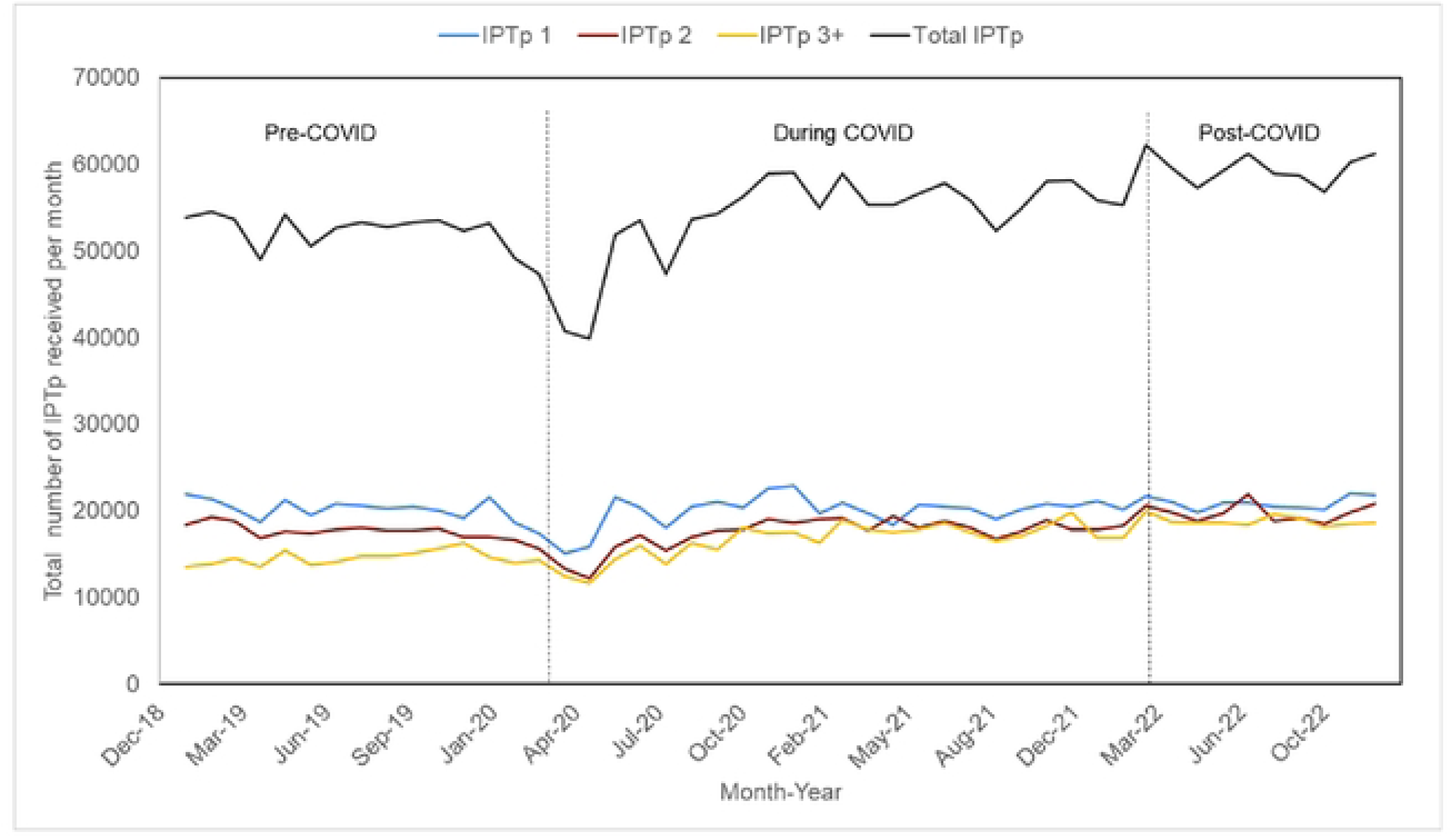
Total IPTp and IPTp 1,2 and 3+ receipt per month in Sierra Leone, over three time periods. IPTp (Intermittent Preventive Therapy).

IPTp1 uptake is highest in all the periods, with lower IPTp2 and then IPTp3+ uptake across all periods. IPTp2 and IPTp3 both had a more marked increase in uptake following recovery than IPTp1, though all experienced sustained increase after COVID-19 (**Figure 1)**.

### Geographic variation in the uptake of IPTp before, during and after COVID-19 across the five regions of Sierra Leone

We disaggregated the data by the five regions (East, North, North-West, South, and Western Area) of Sierra Leone to look for regional variation in trends. All regions had stable fluctuations through all time periods and all regions had a similar decline in uptake starting in January 2020, the last three months of the pre-COVID-19 period. Some regions had a sharp increase in total uptake in early 2021, notably the Northern and Eastern regions and the Eastern region had a slower recovery after the decline compared to the other regions **(Figure 2).**

**Figure 2:**
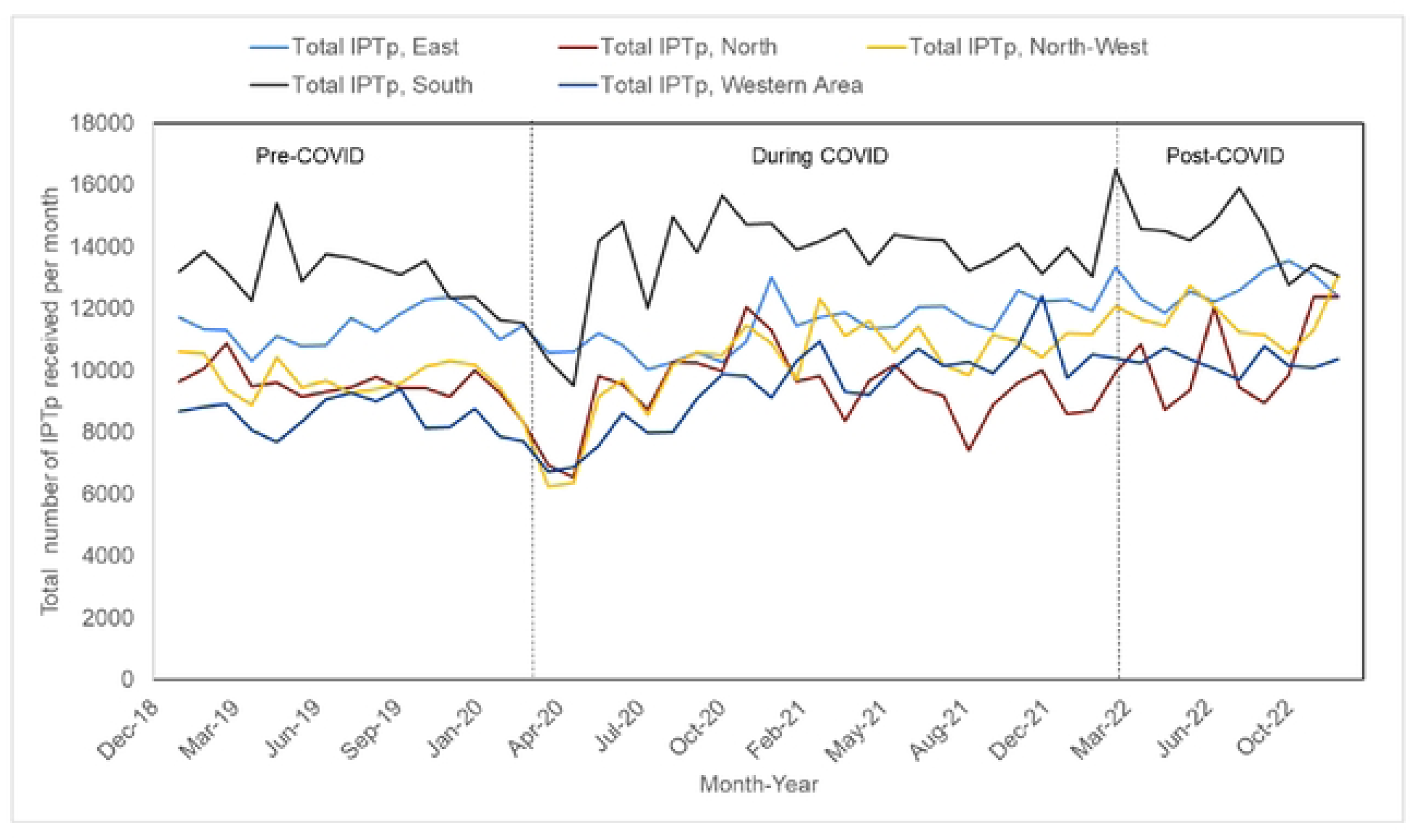
Total IPTp (intermittent preventive therapy) during pregnancy, received per month over three time periods in the five regions in Sierra Leone.

## Discussion

Using longitudinal aggregate program data, we found a dynamic pattern in the uptake of IPTp over three time periods shaped by the COVID-19 pandemic in Sierra Leone. We observed small fluctuations in the IPTp-SP uptake throughout the three time periods of COVID-19; a decline in uptake of IPTp in the latter part of the pre-COVID-19 period with further decline during the early phase of the COVID-19 pandemic; and a delay in recovery of IPTp uptake in the Eastern region compared to the other four regions of Sierra Leone.

The fluctuations in IPTp throughout all the three time periods could be related to a number of factors that include management of the supply of SP (12), issues of stock-out (13) throughout the three time periods in this study, and seasonal variations of the COVID-19 cases. Despite the challenges in the availability of resources, and the occasional issues of stock-out, the program was able to provide a continuous supply of SP for all three IPTp types until the post-COVID-19 period. This was possibly because of the increased programmatic awareness of the challenges affecting the prevention of malaria in pregnancy and especially with the uptake of IPTp uptake (14). A qualitative study in Uganda on the barriers to uptake of IPTp revealed that supply side chain management and refusal of SP account for most of the missed IPTp doses in antenatal care and suggests the need for IPTp safety reassurance among pregnant women(15).

Overall, the most notable finding was a very clear decline in uptake of IPTp that started in January 2020 that had its nadir prior to the national lockdowns in April 2020, followed by recovery by mid-2020, and a further increase by early 2021 and into the post-COVID-19 period. This finding was in the background of an otherwise relatively steady increase over time of uptake of IPTp across all periods and was similar across all geographical regions, except the Eastern region which had a slower recovery. The decline in services could have started several months before the country’s first case (31 March 2020) and prior to the implementation of lockdown and intercountry travel restriction in April 2020. Preparation for the pandemic began in late 2019 to pre-emptively address population anxiety and community fear rooted in the impact of the 2014-2016 Ebola outbreak in Sierra Leone(16)(17). Government mitigations in that period included shifting resources away from all ministries, departments, and agencies to address the needs of the COVID-19 preparedness plan with reports suggesting possible effects on community health worker motivation, low levels of essential drugs in particular at the PHU levels, and limited supply drugs for use at the facilities and communities (18). A systematic review in sub-Saharan Africa confirmed that some of the barriers to improving IPTp uptake are poor leadership and governance, low budgetary allocation, low staff motivation, long distances and long waiting times, drug stock outs, and poor management of information and supply chain. Therefore, the need for malaria control programs in Africa to address the barriers to low coverage and program ineffectiveness and prioritize increase in the uptake of IPTp to prevent malaria in pregnancy (19).

Across all periods, IPTp1 uptake was highest with lower uptake of IPTp2 and then IPTp3+ across all periods with the same pattern observed amongst IPTp1, 2, and 3+ regarding the timing and magnitude of decline and recovery during COVID-19. The recovery and increase IPTp uptake during COVID-19 and post COVID-19 appeared to be more marked with IPTp2 and 3+ such that levels converged in the post-COVID-19 period. The recovery phase of IPTp uptake showed a steady increase, which was maintained post-COVID-19. This overall trend of improvement in IPTp indicators, particularly of IPTp2 and IPTp3+ across the time periods, reflects some programmatic resiliencies that had just started before COVID-19.

Of note, in October 2019, Impact Malaria, a USAID initiative(20) was launched to support the treatment and prevention of malaria in pregnancy. Commencement of training for healthcare workers (including community health workers) on malaria in pregnancy (IPTp uptake and other related malaria services) started in October 2020. The slower recovery in the Eastern region could partly be explained by the delay in rolling-out of the malaria in pregnancy initiative in this region, which started in the North and Southern regions before expanding to the Eastern region. Ongoing support to the Community Health Worker initiative continued throughout the study period by the Global Alliance Vaccine Initiative in Kono, Karene, and Bonthe and the U.S. President’s Malaria Initiative (Sierra Leone) in Kailahun, Falaba, and Pujehun (21).

Our study suggests the need to provide appropriate measures to address supply chain issues, and safety reassurance on the use of IPTp for the prevention of malaria among pregnant women. There is also the need to have a system in place at national, regional, and district levels that addresses emergency preparedness and readiness in disease outbreaks using a risk management approach to ensure that essential services continue uninterrupted.

The strength of the study is that the use of data from district-level programme information collected as part of routine monitoring and evaluation from a national public system should be representative of the country. Study limitations are the magnitude of data computation required, and the reliance on the human element for data transfer, input, and data abstraction that could impact the data in the DHIS-2 platform that was used for this analysis.

## Conclusion

There has been progress with IPTp uptake that created some resiliency in services during the COVID-19 pandemic so that incremental progress could continue. Nonetheless, a critical piece of pandemic preparedness and early response should incorporate planning so that ongoing services, including those at the community level, are continued without interruption. Ongoing technical resource assistance through programs like Impact Malaria was an effective strategy in the medium term for creating resiliency in services addressing malaria in pregnancy.

## Declaration section

The authors declared no conflict of interest

## Acknowledgements

We acknowledge the staff and members of National Malaria Control Program of the Ministry of Health for providing the enabling environment to conduct this study. We thank Kristy Yiu (Unity Health Toronto) for her early support in coordination of the project. Finally, we thank the staff of Sustainable Health System for their support. SM is supported by a Tier 2 Canada Research Chair in Mathematical Modeling and Program Science.

## Author’s contribution

Conceptualization and study design: WTP, ML, SL, SS

Data curation and validation: WTP, MSK, BS, WKL, AMF

Methodology and formal analysis: BM, WTP, ML, MSK, SM

Funding acquisition, resources, supervision: SS, SL, SM, ACK

Writing – original draft preparation: WTP, ML, ACK

Writing – review & editing: MSK, BS, WKL, AMF, SM, SS, SL

## Availability of data

The dataset (https://doi.org/10.5061/dryad.4tmpg4fmx) is available from: https://datadryad.org/stash/dashboard.

